# Cerebrospinal Fluid Pharmacokinetics of Nicardipine Following Intrathecal Administration in Subarachnoid Hemorrhage Patients

**DOI:** 10.1101/2023.10.17.23297116

**Authors:** Ofer Sadan, Yoo-Seong Jeong, Shany Cohen-Sadan, Eashani Sathialingam, Erin M. Buckley, Prem A. Kandiah, Jonathan A. Grossberg, William Asbury, William J Jusko, Owen B. Samuels

## Abstract

Subarachnoid hemorrhage (SAH) is a devastating type of stroke, leading to high mortality and morbidity rates. Cerebral vasospasm and delayed cerebral ischemia (DCI) are common complications following SAH and contribute significantly to the poor outcomes observed in these patients. Intrathecal (IT) nicardipine delivered via an existing external ventricular drain has been shown to be correlated with reduced DCI and improved patient outcomes. The current study aims to characterize population pharmacokinetic (popPK) properties of intermittent IT nicardipine. Following informed consent, serial cerebrospinal fluid (CSF) samples were obtained from 16 SAH patients (50.4 ± 9.3 years old; 12 females) treated with IT nicardipine every 6 hours (n=8) or every 8 hours (n=8), which were subject to high-performance liquid chromatography for measurement of its CSF concentration. Our popPK analysis showed that the CSF PK of IT nicardipine in the cohort was adequately described by a two-compartment model with a lag time, with reliable parameter estimates (relative standard error < 50%). The intracranial pressure influenced both the total clearance and the central volume. Calculated PK parameters were similar between q6h and q8h dosing regimens. Despite a small cohort of SAH patients, we successfully developed a popPK model to describe the nicardipine disposition kinetics in the CSF following IT administration. These findings may help inform future clinical trials designed to examine the optimal dosing of IT nicardipine.

## INTRODUCTION

Subarachnoid hemorrhage (SAH) is a devastating type of stroke, leading to high mortality and morbidity rates. Despite advancements in care, the in-hospital mortality rate for SAH patients remains high,^1,2^ and even survivors often have limited functional outcomes with many unable to return to their previous occupation.^3^ Cerebral vasospasm and delayed cerebral ischemia (DCI) are common complications following SAH and contribute significantly to the poor outcomes observed in these patients.^4^ The relationship between cerebral vasospasm and DCI remains unclear, yet the development of vasospasm is known to be a risk factor for the development of DCI later in the course.^5–7^ Unfortunately, effective treatment strategies to address cerebral vasospasm and prevent DCI are lacking. Current guidelines recommend the use of prophylactic oral nimodipine to prevent DCI,^8^ however, no other high-quality data-based interventions are available when cerebral vasospasm and/or DCI develops.

Decades of searching for interventions to mitigate these complications following SAH have not resulted in a clear positive effect on patient outcomes, even in well-powered phase III trials. This is despite multiple attempts using different therapeutic strategies.^9^ One example was the use of intravenous (IV) nicardipine, a dihydropyridine drug similar to nimodipine, administered via continuous infusion. The prophylactic IV infusion of nicardipine administered to patients with SAH did reduce the incidence of cerebral vasospasm, however, it did not change patient outcomes.^10^ This result likely stems from hypotension induced by this mode of nicardipine administration. Hypotension, in the setting of cerebral vasospasm, reduces cerebral perfusion and may offset the positive vasodilator effect of nicardipine. Another potential reason for the reduced efficacy of IV nicardipine in this population could be due to its low brain-to-systemic concentration ratio at usual infusion rates (2.5-15 mg/hour), despite its known hydrophobicity (log P = 3.82).^11^ In a rat model, a low brain tissue exposure has been indeed observed when nicardipine was orally administered.^12^ Therefore, systemic administration of nicardipine may require higher doses to achieve its sufficient efficacy in the central nervous system (CNS), which are more likely to result in hypotension.

Given the limitation of IV nicardipine in preventing or treating cerebral vasospasm or DCI, direct administration of parenteral nicardipine into the brain has been proposed as an option. Intrathecal (IT) administration is now a commonly utilized route for drug delivery,^13,14^ since methotrexate was first given IT to CNS leukemia patients.^15^ Although the IT route may pose specific risks such as increased intracranial pressure (ICP), CNS infection or other toxicity,^16^ this route has several therapeutic advantages, especially when (i) the therapeutic target is present within the subarachnoid space or CSF compartment (e.g., meninges, blood vessels), (ii) the drug molecules in the systemic circulation do not have sufficient access to the spinal space (e.g., for hydrophilic drugs and large molecules such as antibodies and oligonucleotides), and/or (iii) the off-target toxicity of the therapeutic agent needs to be avoided. For instance, baclofen, which is a hydrophilic drug used for treatment of spinal cord spasticity, is utilized at a maximal IT dose of 0.1 mg/day, while the oral dose may reach 120 mg/day.^17^ Typically, the volume of distribution of IT administered drugs is expected to be relatively small (∼ 80 mL; the spinal CSF space or ∼150mL when considering the brain compartment as well),^18^ and the CSF half-life for IT administered drugs appears to be consistently short, ranging from 1.2 to 2.7 hours.^14^

Given the reported efficacy of orally administered calcium channel blockade following SAH, IT administration of nicardipine is an attractive consideration that should provide high brain concentrations while minimizing systemic exposure. This has led to a widening adoption of IT nicardipine delivered via an external ventricular drain (EVD). Growing evidence, albeit mostly uncontrolled and retrospective, suggests that this treatment is safe and associated with reduced rates of DCI and poor functional outcomes.^5,19^ A quantitative assessment of nicardipine disposition in the CSF following IT administration has not been done. Thus, this report describes the creation of a population pharmacokinetic (popPK) model based on nicardipine concentration-time profiles in the CSF following repeated IT dosing in 16 SAH patients.

## METHODS

### Patient population

Patients who were admitted with a non-traumatic SAH, required an EVD, and later were treated with IT nicardipine for cerebral vasospasm were recruited for this study. This observational study enrolled patients admitted to the Emory University Hospital (Atlanta, GA, USA) neuroscience intensive care unit (neuroICU), following approval by the Emory University institutional review board (IRB00107474) and signed consent by the patient or their legally authorized representative. Patient demographics, cardiovascular risk factors, and clinical course were recorded. All data produced in the present study are available upon reasonable request to the authors.

### Data collection

The CSF concentration-time data for IT nicardipine were obtained from 16 patients. Briefly, all the 16 patients were given a bolus injection of 5 mg nicardipine (2.5 mg/mL as a hydrochloride form; Exela Pharma Sciences, Lenoir, NC, USA) via IT route (over 1-2 minutes) every 6 hours (n=8) or 8 hours (n=8) as part of usual care. Our usual starting dose of IT nicardipine was 5 mg every 8 hours, and the dose and frequency of administration were at the discretion of the ICU attending physician. The decision for the specific regimen was made by the clinical team and was not protocolized. The CSF was collected via the collection chamber in the EVD system every hour up to 6 or 8 hr (i.e., depending on the dosing interval) after the first dose and another dose on the third day of treatment (e.g., dose #6-9). The EVD system that was used for this study is graphically described in Supplementary Figure S1.

### HPLC measurements

The CSF samples (100 μL) were vortex-mixed with an equal volume of methanol, and the resulting mixtures were placed in ice for at least 30 minutes. The samples were subsequently centrifuged at 4°C for 15 minutes at 13,000 rounds per minute, and the supernatant was diluted with an equal volume of double distilled water. The 50 μL of the diluted sample was injected for HPLC (Waters, Milford, MA, 1525 binary pump) assay equipped with photodiode array autosampler (Waters model 2996). Analytes were separated using reverse-phase chromatography on a Waters Xbridge BEH column (3×150 mm, 3.5 µM, Waters) maintained at 4°C. The mobile phase consisted of (A) 10 mM trimethylamine at pH 3.5 (adjusted with phosphoric acid) and (B) acetonitrile (i.e., an isocratic condition at A:B = 65:35). The flow rate of the mobile phase was 0.5 mL/minute, and the run time was 20 minutes. The chromatograms were monitored at a wavelength of 240 nm.

### PopPK model development

The popPK model was developed using a stepwise approach. The structural model that best described nicardipine concentration-time relationships in the CSF was selected based on all patient data. One- and two-compartment models with and without a lag time were tested for a base model. The inter-individual variability (IIV) was first added to each model parameter and only removed when the IIV was very small with a high relative standard error (RSE). Parameters were estimated using the method of stochastic approximation of expectation maximization (SAEM)^20^ built in Monolix 2023R1 (Lixoft SAS; a Simulations Plus company).

For all covariates, the baseline value was defined as the first measurement of the study period, which is summarized in Table 1. In the case of stationary covariates [e.g., age, body weight, height, body surface area (BSA), sex, race, initial World Federation of Neurosurgical Societies (WFNS) scale, modified Fisher grade (mFG); Table 1], candidate covariates were identified based on assessment of their impact on the specified base model parameters: A covariate was subject to covariate searching when a statistical significance (*p* < 0.05) was found with the specified base model parameter, based on Pearson’s correlation test (continuous covariates) or analysis of variance (categorical covariates). The appropriateness of the covariate included in the model was post-hoc tested for significance using Wald’s test.^21^ All time-varying covariates [e.g., Q_CSF_, intracranial pressure (ICP), systolic blood pressure (SBP), diastolic blood pressure (DBP), heart rate (HR), and O_2_ saturation (SpO_2_); Table 1] were also included in the list of candidate covariates.

**Table 1.** Patient characteristics and covariate summary.

| Variable at baseline (unit) | Minimum | First<br>quartile | Median | Mean | Third<br>quartile | Maximum |
| --- | --- | --- | --- | --- | --- | --- |
| Age (years) | 37 | 43.8 | 50.4 | 49 | 58 | 65 |
| Body weight (WT; kg) | 56.3 | 69.7 | 82.6 | 74.8 | 88.5 | 142 |
| Height (HT; cm) | 145 | 160 | 165 | 165 | 173 | 182 |
| Body surface area (BSA; m<sup>2</sup>) | 1.63 | 1.69 | 1.88 | 1.79 | 2.01 | 2.48 |
| **Categorical variables** | **Group (size)** |  |  |  |  |  |
| Sex | Male (3), female (13) |  |  |  |  |  |
| Race/ethnicity | African American (6), White (7), Unknown (3) |  |  |  |  |  |
| WFNS grade | Grade 1 (0), Grade 2 (8), Grade 3 (0), Grade 4 (5), Grade 5 (3) |  |  |  |  |  |
| Modified Fisher grade (mFG) | Grade 1 (0), Grade 2 (0), Grade 3 (4), Grade 4 (12), Grade 5 (0) |  |  |  |  |  |
| Value at baseline (unit) | Minimum | First<br>quartile | Median | Mean | Third<br>quartile | Maximum |
| Output flow of CSF ($Q_{CSF}$; mL/hr) | 0 | 5 | 8.56 | 9 | 12 | 17 |
| Intracranial pressure (ICP; mmHg) | 2 | 6.5 | 8 | 8 | 10 | 16 |
| Systolic blood pressure (SBP; mmHg) | 118 | 136 | 152 | 148 | 163 | 196 |
| Diastolic blood pressure (DBP; mmHg) | 55 | 70 | 76.8 | 76 | 82.3 | 108 |
| Heart rate (HR; bpm) | 49 | 68.8 | 79.3 | 78 | 89 | 107 |
| O<sub>2</sub> saturation (SpO<sub>2</sub>; %) | 93 | 96 | 97.6 | 98 | 99 | 100 |

A univariate method was used for covariate searching under the criteria of *p* < 0.01 for forward inclusion and *p* < 0.001 for backward elimination, based on the objective function values (OFV). Continuous covariate (x_i_)-parameter (P_i_) relationships were examined using a power model (Eq. 1):

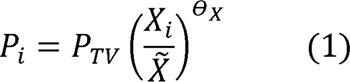

where e_X_ is the power estimate for the covariate effect, P_TV_ is the typical value of the parameter, and x is the median value of the covariate. In addition, categorical covariate (r_i_)-parameter (P_i_) terms were modeled using Eq. 2:

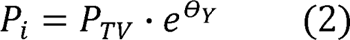

where e_Y_ is the covariate effect parameter in terms of r. Covariate searching was conducted using an automatic covariate search function in Monolix software where all stationary covariate-parameter relationships are tested. The statistical significance of implementing a time-varying covariate on a specific model parameter was also tested by considering the time-varying covariate as a regressor in Monolix.

The final popPK model underwent graphical and numerical goodness-of-fit (GOF) analyses as well as prediction-corrected visual predictive checks (VPCs). During the graphical GOF analysis, diagnostic plots were carefully examined, plotting observed versus predicted values, to identify any indications of systematic lack of fit or bias in the error distributions. The numerical GOF assessment utilized several criteria, such as ensuring successful numerical convergence, acceptable parameter precision (with a relative standard error < 50%), a low condition number (< 100) to maintain model stability, and biological plausibility of the PK parameter values based on existing knowledge of the drug’s behavior.

## RESULTS

### Patient cohort

Table 1 provides the summary of demographic and clinical characteristics for 16 patients who were hospitalized with SAH. The study cohort consisted of 12 (81%) females, with an average (± standard deviation) age of 50.4 ± 9.3 years. The mean body weight was 82.6 ± 22.0 kg; height was 165 ± 9.13 cm; and BSA was 1.89 ± 0.238 m^2^. Of note, there was no statistically significant difference between the groups of patients who were treated with a q6h regimen and a q8h regimen in regard to the demographics, risk factors, or bleed severity (data not shown). This study provides 117 dosing records and 256 concentrations. There were 26 data points found below the quantitation limit (i.e., 50 ng/mL), which were thus assumed to fall between 0 and 50 ng/mL.^22^ These observations were subsequently used for the popPK modeling.

### Population PK analysis

The CSF PK of IT nicardipine was adequately described by a two-compartment model with a lag time (see Figure 1 showing the model scheme and differential equations). The following parameters of the structural PK model were estimated: Central volume (v_1_), peripheral volume (v_2_), total clearance (CL), inter-compartmental clearance (CL_D_), and lag time (r_lag_). The IIV in v_1_, v_2_, CL, and r_lag_ was incorporated into the structural model. The individual fitting results are provided in Figure 2, and the basic goodness-of-fit plots of the final PK model are given in Supplementary Figure S2. The conditional weighted residual plots did not show any trends, all ranging within ± 4.

**Figure 1.**
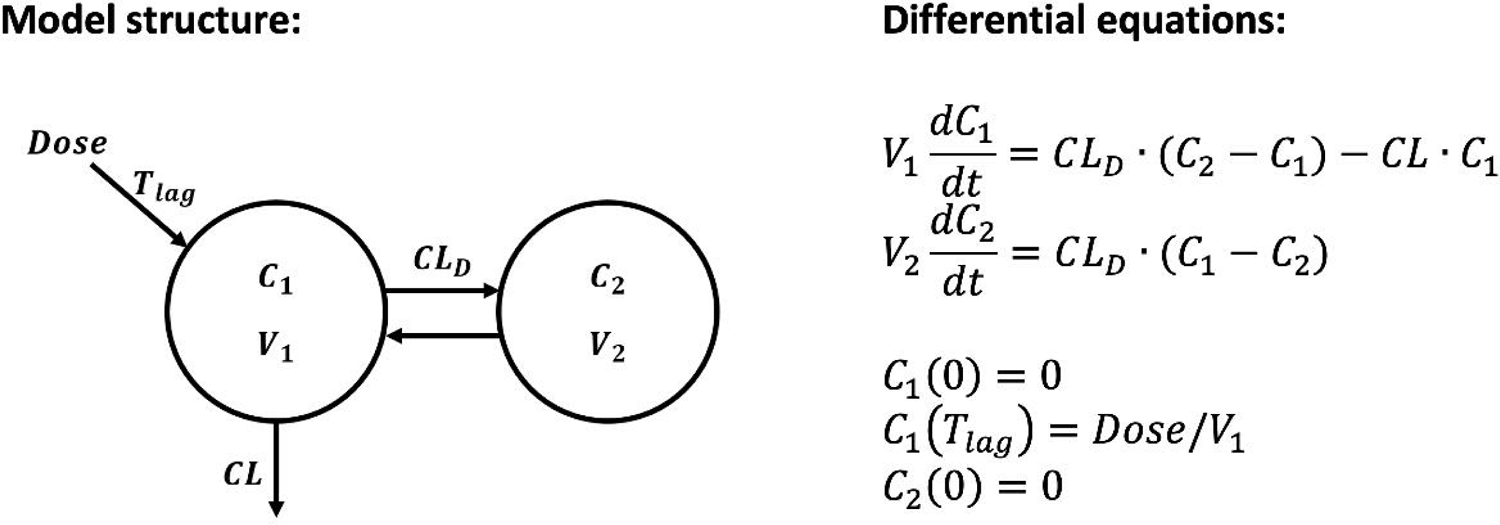
Model structure and differential equations used for development of a popPK model for IT nicardipine. Symbols and definitions are provided in Table 2.

**Figure 2.**
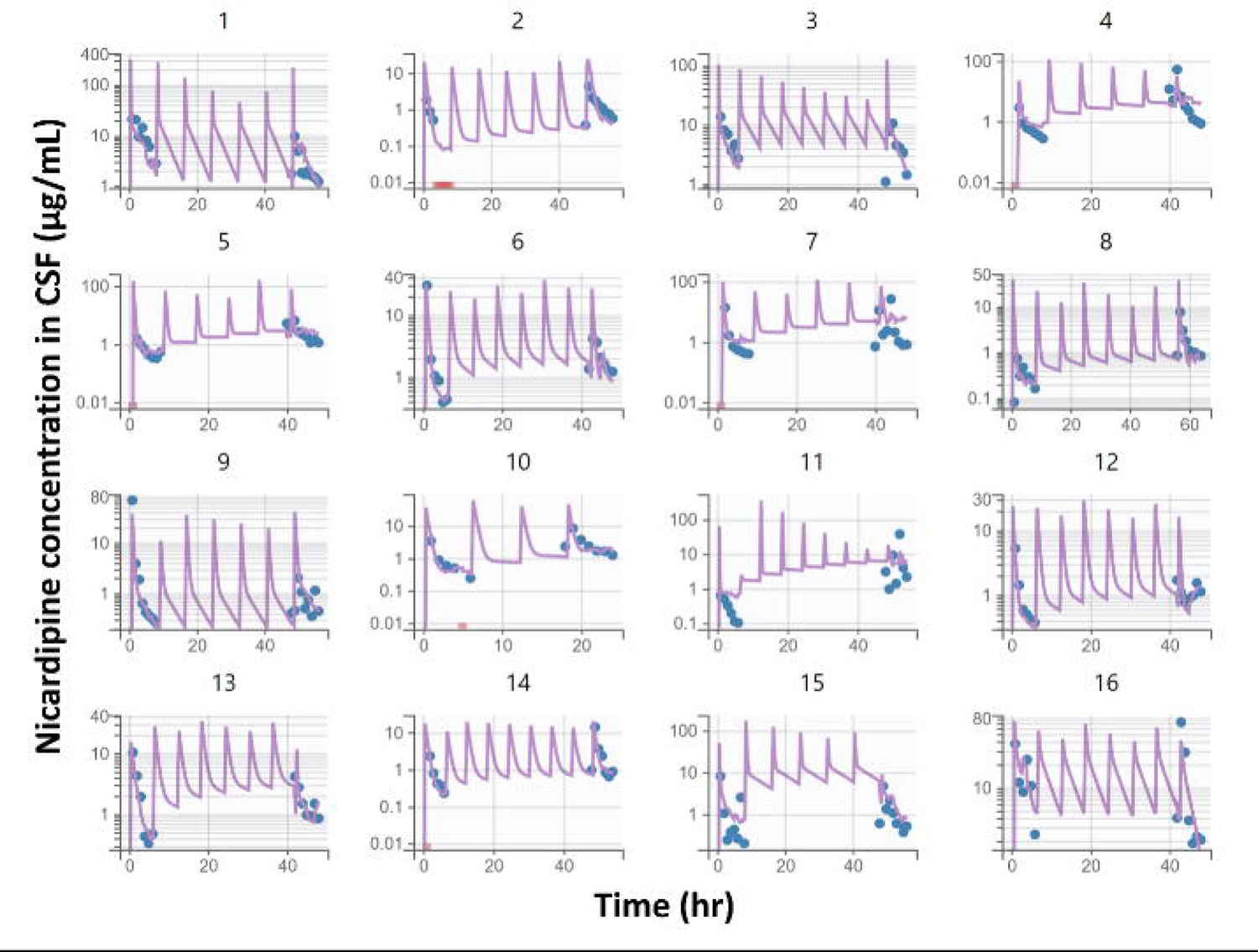
Individual fittings of the CSF concentration versus time profiles for nicardipine following its IT doses of 5 mg to 16 patients with SAH. Blue dots are measured concentrations and purple lines are model-fitted profiles for each individual; pink dots are the data below the lower limit of quantification (50 ng/mL). Serial patient ID numbers are indicated.

**Table 2.** Parameter estimates from the final popPK model for IT nicardipine in SAH patients (%) relative standard error (RSE), [%] shrinkage.

| Parameter (unit) | Definition | Estimate | Inter-individual variability (IIV) |
| --- | --- | --- | --- |
| $V_1$ (L) | Central volume | 0.064 (35.5) | 1.04 (27.2) [21.3%] |
| $CL$ (L/hr) | Total clearance | 0.130 (19.8) | 0.69 (23.1) [8.83%] |
| $V_2$ (L) | Peripheral volume | 0.73 (41.7) | 1.38 (21.1) [2.86%] |
| $CL_D$ (L/hr) | Inter-compartmental clearance | 0.11 (2.06) | - |
| $T_{lag}$ (hr) | Lag time | 0.55 (20.2) | 0.6 (27.8) [29.5%] |
| $\theta_{ICP,CL}$ | ICP effect on $CL$ | 1.46 (3.47) | - |
| $\theta_{ICP,V1}$ | ICP effect on $V_1$ | 0.35 (0.899) | - |
| Residual variability |  |  |  |
| Proportional error |  | 0.69 (6.61) |  |

The experimental data and fittings shown in Figure 2 exhibit bi-exponential profiles consistent with the initial distribution into brain tissue and the overall loss out of the CSF. The first- and last-dose profiles show a modest accumulation of nicardipine owing to repeated dosing and the linear and stationary PK as assumed in the modeling.

### Covariate analysis and model evaluation

We assessed the impact of candidate covariates on the structural parameters, i.e., age (on r_lag_), and Q_CSF_, ICP, SBP, DBP, HR, and SpO_2_ (on r_lag_, CL, v_1_, and v_2_). Although SBP seemed to be a statistically significant covariate for r_lag_ (ΔOFV = −13.49, *p* < 0.001), another significant covariate ICP on CL (ΔOFV = −12.52, *p* < 0.001) was chosen for further covariate searching. Subsequently, the ICP was a significant covariate also for v_1_ (ΔOFV = −20.82, *p* < 0.001). When we focused on the time-zero kinetic properties (i.e., the CL and v_1_ values calculated by using the baseline ICP value), a significant correlation was found between the ICP and the CL (R^2^ = 0.5965), while the individual v_1_ values at time 0 were not enough to represent a statistical significance (R^2^ = 0.07857) (Figure 3A). Since the ICP was found to be related to both CL and v as shown in Figure 3B, we also attempted to include the correlation between CL and v_1_ in the model. However, inclusion of this CL versus v_1_ correlation in a separate fitting process did not improve the model performance (ΔOFV = +4.23 or +22.1). The parameter estimates and IIV values of the final model are summarized in Table 2. The VPC plot (Supplementary Figure S3) indicate that the final model adequately encompassed the trend and variability of the observed nicardipine concentrations in the CSF.

**Figure 3.**
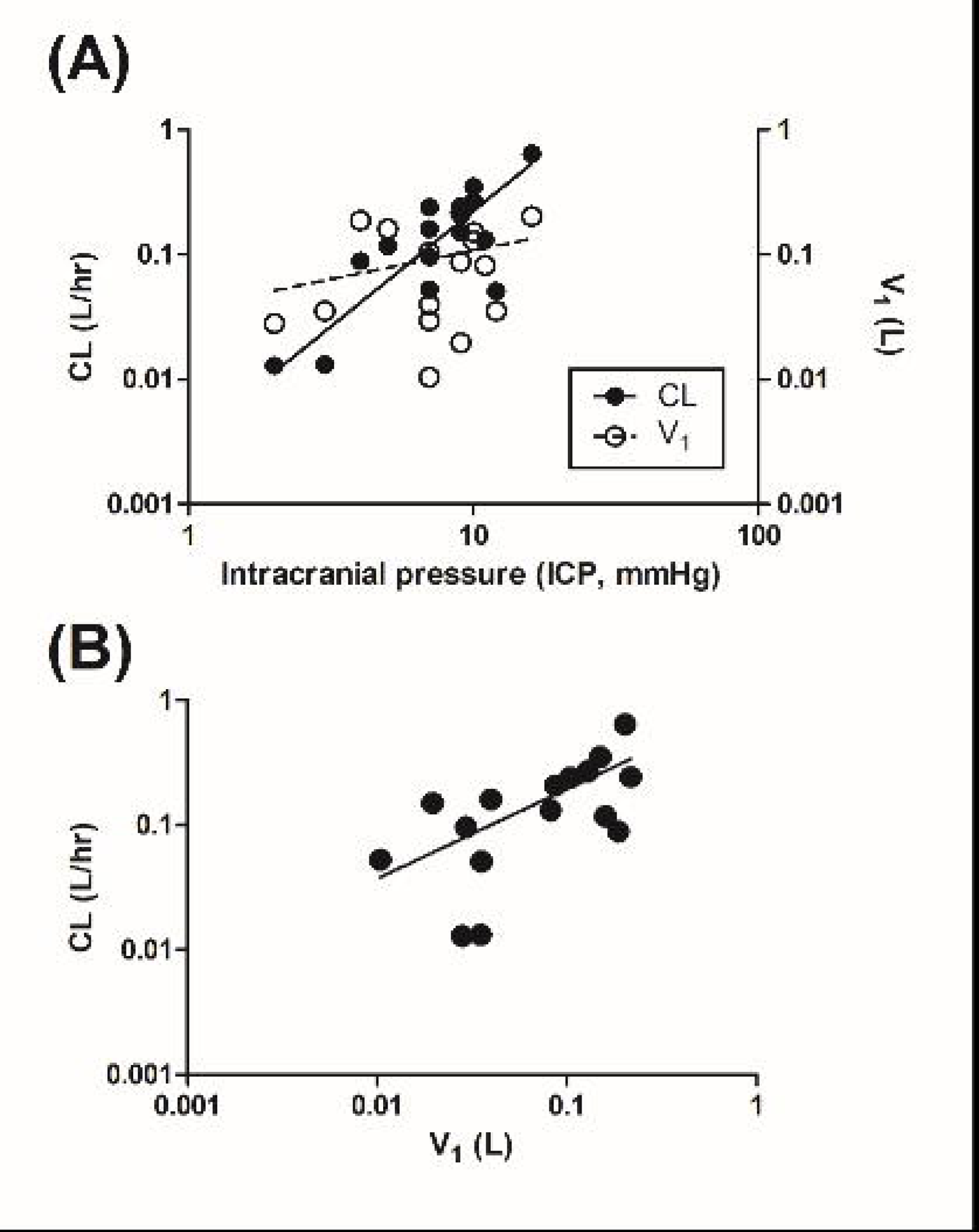
Correlations (A) of the CL (closed circles and solid line) or v_1_ (open circles and dashed line) versus the intracranial pressure (ICP), and (B) between the CL and v_1_ values, based on the final popPK model. All the CL and v_1_ parameters are empirical Bayes estimates at time 0 (i.e., baseline)

## DISCUSSION

Our popPK model for nicardipine PK in the CSF of 16 SAH patients functioned well. Despite the diverse and critically ill state of the patients receiving the drug, the nicardipine concentration-time profiles in the CSF of the patients were reasonably described in a consistent manner by the two-compartment model with a lag time (Figure 2 and Supplementary Figure S3). The RSE values of the parameter estimates (< 50%; Table 2) support the reliability of the current model.

In order to verify that the methods used to collect the CSF samples are valid, we performed an in-vitro control experiment (see supplementary methods). These results confirmed that nicardipine did not significantly adsorb to the EVD or collection system (Supplementary results, Figure S1). Nicardipine concentrations in plasma were also monitored at 0, 0.25, 1, 4, 6, and 8 hours after its injection into the CSF. However, no nicardipine was measured above the lower limit of quantification (50 ng/mL), suggesting that the transfer of nicardipine into the systemic circulation could not be discerned. Selected plasma samples were measured using liquid chromatography-mass spectrometry (LC/MS), which verified that the plasma concentration was below 50ng/mL (data not shown). Considering that its CSF concentrations were found to approximately range from 0.1 to 100 mcg/mL (Figure 2), the apparent drug partitioning into the systemic circulation may be kinetically insignificant, based on the CSF-to-plasma concentration ratios possibly ranging from 2,000 to at least 2. When nicardipine was orally administered (and thus systemically exposed first) to rats,^12^ its exposure to the brain appears limited based on a brain-to-plasma ratio of approximately 0.1. Collectively, therefore, we reasoned that the blood-brain-barrier would block the transport of nicardipine in both directions, i.e., into and out of the brain.

Based on these findings, we focused only on assessment of the nicardipine PK within the brain, rather than its systemic effect. It can be noted that the v_1_ of 0.064 L is comparable to the physiological volume of the CSF (e.g., 25 mL in the ventricles and 125 mL in the subarachnoid spaces),^23^ and the v of 0.73 L is around 50% of the average brain weight of 2% of body weight.^24^ The numerical difference between the inter-compartmental clearance (0.11 L/hr; Table 2) and the brain extracellular fluids (ECF, 0.0105 L/hr) and CSF (0.0240 L/hr) flow^25^ implies that, in addition to the interstitial fluid, there could be more distributional components within the brain mass.

The central volume and the total clearance of nicardipine was found to correlate with the ICP. Based on the e_lCP,V1_ value (0.35; which is larger than 0; Table 2), our finding is consistent with the intracranial volume-pressure relationship that correlates positively.^26,27^ Of note, the baseline relationship between the ICP and v_1_ parameters (Figure 3A) was not sufficient to represent their time-dependent correlations. While the inclusion of the correlation between CL and v_1_ into the statistical model structure did not improve the model performance (see Results), the higher ICP is expected to also increase the nicardipine clearance from the CSF (e_lCP,CL_ value of 1.46; Table 2). As shown in Figure 3A, a statistical significance could be obtained even only with the baseline relationship between the ICP and CL. Interestingly, the quantitative CSF output (Q_CSF_) was not correlated to the clearance. The combination of these findings suggests that the elimination mechanism of nicardipine out of the CSF may include its transfer into the physiological sink, rather than its permanent elimination via the EVD. The elimination pathway is likely affected by fluid pressure within the ventricles, which stems from the current volume-pressure curve relationship (i.e., intracranial compliance).

This study possesses some limitations. Although distribution of the individual model parameters could be determined quite reliably (e.g., shrinkage < 30%; Table 2) with the limited sample size (n = 16), further study is warranted to develop a more comprehensive population model. With additional PK and patient outcome data perhaps a more optimal IT dosing strategy can evolve.

## CONCLUSIONS

With a small cohort of SAH patients, we successfully developed a popPK model to describe the nicardipine disposition kinetics within the CSF following direct IT administration. CSF concentration-time profiles of nicardipine were reasonably described using a two-compartment model with a lag time. The nicardipine PK characteristics in the CSF were similar between q6h and q8h dosing regimens. The ICP was noted to be a significant covariate for both the central compartment volume of distribution (v_1_) and total clearance (CL). Our findings may be helpful in designing subsequent clinical trials to further evaluate the PK of nicardipine in the CSF, and to ultimately determine optimal nicardipine dosing strategies that will lead to improved outcomes for SAH patients.

## Data Availability

All data produced in the present study are available upon reasonable request to the authors

## Acknowledgements

This study was supported in part by the National Institutes of Health under Award Number R01NS13003601 (OS). It was also supported in part by the Emory HPLC Bioanalytical Core (EHBC), which was supported by the Department of Pharmacology, Emory University School of Medicine and the Georgia Clinical & Translational Science Alliance of the National Institutes of Health under Award Number UL1TR002378. Drs. Jeong and Jusko were supported by NIH grant R35 GM131800. The content is solely the responsibility of the authors and does not necessarily reflect the official views of the National Institutes of Health.

EVDs for the in-vitro experiment were contributed by the manufacturer (Codman). The supplier did not intervene in study design, analysis or writing of the manuscript.

## REFERENCES

1. Udy AA, Vladic C, Saxby ER, et al. Subarachnoid Hemorrhage Patients Admitted to Intensive Care in Australia and New Zealand: A Multicenter Cohort Analysis of In-Hospital Mortality Over 15 Years. Critical care medicine. 2017;45(2):e138–e145. doi:10.1097/CCM.0000000000002059

2. Samuels OB, Sadan O, Feng C, et al. Aneurysmal Subarachnoid Hemorrhage: Trends, Outcomes, and Predictions From a 15-Year Perspective of a Single Neurocritical Care Unit. Neurosurgery. 2021;88(3):574–583. doi:10.1093/neuros/nyaa465

3. Al-Khindi T, Macdonald RL, Schweizer TA. Cognitive and functional outcome after aneurysmal subarachnoid hemorrhage. Stroke. 2010;41(8):e519–36. doi:10.1161/STROKEAHA.110.581975

4. Pegoli M, Mandrekar J, Rabinstein AA, Lanzino G. Predictors of excellent functional outcome in aneurysmal subarachnoid hemorrhage. Journal of neurosurgery. 2015;122(2):414–418. doi:10.3171/2014.10.JNS14290

5. Sadan O, Waddel H, Moore R, et al. Does intrathecal nicardipine for cerebral vasospasm following subarachnoid hemorrhage correlate with reduced delayed cerebral ischemia? A retrospective propensity score–based analysis. Journal of Neurosurgery. 2021;136(1):115–124. doi:10.3171/2020.12.JNS203673

6. Snider SB, Migdady I, LaRose SL, et al. Transcranial-Doppler-Measured Vasospasm Severity is Associated with Delayed Cerebral Infarction After Subarachnoid Hemorrhage. Neurocritical care. Published online November 9, 2021:1–7. doi:10.1007/s12028-021-01382-2

7. Frontera JA, Fernandez A, Schmidt JM, et al. Defining vasospasm after subarachnoid hemorrhage: what is the most clinically relevant definition? Stroke. 2009;40(6):1963–1968. doi:10.1161/STROKEAHA.108.544700

8. Hoh BL, Ko NU, Amin-Hanjani S, et al. 2023 Guideline for the Management of Patients With Aneurysmal Subarachnoid Hemorrhage: A Guideline From the American Heart Association/American Stroke Association. Stroke. Published online May 22, 2023:STR.0000000000000436. doi:10.1161/STR.0000000000000436

9. Qureshi AI, Lobanova I, Huang W, et al. Lessons Learned from Phase II and Phase III Trials Investigating Therapeutic Agents for Cerebral Ischemia Associated with Aneurysmal Subarachnoid Hemorrhage. Neurocritical care. 2022;36(2):662–681. doi:10.1007/s12028-021-01372-4

10. Haley EC, Kassell NF, Torner JC. A randomized controlled trial of high-dose intravenous nicardipine in aneurysmal subarachnoid hemorrhage. A report of the Cooperative Aneurysm Study. Journal of neurosurgery. 1993;78(4):537–547. doi:10.3171/jns.1993.78.4.0537

11. Sangster J. LOGKOW A Databank of Evaluated Octanol-Water Partition Coefficients. In:; 1997. Accessed September 18, 2023. https://www.semanticscholar.org/paper/LOGKOW-A-Databank-of-Evaluated-Octanol-Water-Sangster/f1be9b21c94e498b499cfe254dfd794d95f3d358

12. Higuchi S, Sasaki H, Seki T. Pharmacokinetic studies on nicardipine hydrochloride, a new vasodilator, after repeated administration to rats, dogs and humans. Xenobiotica. 1980;10(12):897–903. doi:10.3109/00498258009033823

13. De Andres J, Hayek S, Perruchoud C, et al. Intrathecal Drug Delivery: Advances and Applications in the Management of Chronic Pain Patient. Front Pain Res (Lausanne). 2022;3:900566. doi:10.3389/fpain.2022.900566

14. Hayek SM, Hanes MC. Intrathecal Therapy for Chronic Pain: Current Trends and Future Needs. Curr Pain Headache Rep. 2013;18(1):388. doi:10.1007/s11916-013-0388-x

15. Shapiro WR, Young DF, Mehta BM. Methotrexate: Distribution in Cerebrospinal Fluid after Intravenous, Ventricular and Lumbar Injections. New England Journal of Medicine. 1975;293(4):161–166. doi:10.1056/NEJM197507242930402

16. Kroin JS. Intrathecal drug administration. Present use and future trends. Clin Pharmacokinet. 1992;22(5):319–326. doi:10.2165/00003088-199222050-00001

17. Plassat R, Perrouin Verbe B, Menei P, Menegalli D, Mathé JF, Richard I. Treatment of spasticity with intrathecal Baclofen administration: long-term follow-up, review of 40 patients. Spinal Cord. 2004;42(12):686–693. doi:10.1038/sj.sc.3101647

18. Heetla HW, Proost JH, Molmans BH, Staal MJ, van Laar T. A pharmacokinetic– pharmacodynamic model for intrathecal baclofen in patients with severe spasticity. Br J Clin Pharmacol. 2016;81(1):101–112. doi:10.1111/bcp.12781

19. Hafeez S, Grandhi R. Systematic Review of Intrathecal Nicardipine for the Treatment of Cerebral Vasospasm in Aneurysmal Subarachnoid Hemorrhage. Neurocritical care. 2019;31(2):399–405. doi:10.1007/s12028-018-0659-9

20. Kuhn E, Lavielle M. Maximum likelihood estimation in nonlinear mixed effects models. Computational Statistics & Data Analysis. 2005;49(4):1020–1038. doi:10.1016/j.csda.2004.07.002

21. Harrell, FE. Regression Modeling Strategies: With Applications to Linear Models, Logistic and Ordinal Regression, and Survival Analysis. Springer International Publishing; 2015. doi:10.1007/978-3-319-19425-7

22. Beal SL. Ways to fit a PK model with some data below the quantification limit. J Pharmacokinet Pharmacodyn. 2001;28(5):481–504. doi:10.1023/a:1012299115260

23. L S, G C, J C. Anatomy and physiology of cerebrospinal fluid. European annals of otorhinolaryngology, head and neck diseases. 2011;128(6). doi:10.1016/j.anorl.2011.03.002

24. Davies B, Morris T. Physiological parameters in laboratory animals and humans. Pharm Res. 1993;10(7):1093–1095. doi:10.1023/a:1018943613122

25. Bloomingdale P, Bakshi S, Maass C, et al. Minimal brain PBPK model to support the preclinical and clinical development of antibody therapeutics for CNS diseases. J Pharmacokinet Pharmacodyn. 2021;48(6):861–871. doi:10.1007/s10928-021-09776-7

26. Hawthorne C, Piper I. Monitoring of intracranial pressure in patients with traumatic brain injury. Front Neurol. 2014;5:121. doi:10.3389/fneur.2014.00121

27. Canac N, Jalaleddini K, Thorpe SG, Thibeault CM, Hamilton RB. Review: pathophysiology of intracranial hypertension and noninvasive intracranial pressure monitoring. Fluids Barriers CNS. 2020;17(1):40. doi:10.1186/s12987-020-00201-8

28. Bergstrand M, Hooker AC, Wallin JE, Karlsson MO. Prediction-corrected visual predictive checks for diagnosing nonlinear mixed-effects models. AAPS J. 2011;13(2):143–151. doi:10.1208/s12248-011-9255-z

