## Supplemental for "Cerebrospinal Fluid Pharmacokinetics of Nicardipine Following Intrathecal Administration in Subarachnoid Hemorrhage Patients"

**Supplementary methods**

**In vitro experiment**

To determine whether nicardipine concentrations change following injection through an EVD system, we performed an in vitro experiment. The EVDs (Codman Bactiseal EVD catheter) and collecting systems were identical to the ones used clinically (Supplementary Figure S1). Each experiment was repeated three times, and each sample was measured in triplicate. These in vitro samples were stored at -80°C until the measurement, similar to the biologic samples.

Briefly, four milliliters of nicardipine hydrochloride (HCl) prepared at 2,500 mcg/mL (Exela Pharma Sciences, Lenoir, NC, USA) were injected via the proximal stopcock and through the EVD into a glass container. Samples were obtained at this initial condition (i.e., time 0, at a total of 1 mL with the expected concentration 2,500 mcg/mL), which was followed by injecting 2 mL NaCl 0.9% through the same port in order to mimic the flush injection that occurs clinically. Samples were obtained again (i.e., the expected concentration of 1,500 mcg/mL), and the entire volume was injected through the collecting system into the collecting chamber, from which in the clinical scenario, CSF samples were obtained. From the collecting chamber, samples were obtained after 15, 30 and 60 minutes, while the EVD system was in a lighted room to mimic conditions in an actual patient room. All the collected samples were analyzed using a high-performance liquid chromatography (HPLC) as described in the main text.

**Supplementary Results**

**In vitro assessment of the potential nicardipine adsorption to the EVD tubing**

The in vitro experiment mimicked the nicardipine and saline flush course through an EVD and its collecting system that were used in the clinical settings. The experiment design and its main results are shown in Supplementary Figure S1. When the nicardipine solution (2,500 mcg/mL as a HCl form) went through the EVD system (i.e., before the saline flushing), the resulting nicardipine concentrations were found to be 2,410 ± 193 mcg/mL (96.3 ± 7.72%). After flushing the EVD with the saline, the diluted concentration (i.e., at a theoretical concentration of 1,500 mcg/mL; 3 mL of dosing solution plus 2 mL saline) was observed to be 1,440 ± 185 mcg/mL (95.8 ± 12.3%). In addition, the nicardipine concentration in the collecting chamber appeared consistent at 15 min (1,620 ± 117 mcg/mL), 30 min (1490 ± 178 mcg/mL), and 60 min (1440 ± 33.1 mcg/mL). Collectively, concentrations measured throughout the different locations and times fell within the 96.3-108% range of the expected values, suggesting that nicardipine remained stable and did not significantly adsorb to the EVD or collection system.

**Figure S1**: Nicardipine does not adsorb to the EVD or collecting system tubing. Nicardipine (2,500 mcg/mL, yellow arrow) was injected via the EVD, followed by a NaCl 0.9% flush (blue arrow). The diluted nicardipine (green arrow) was injected back through the EVD collecting system to the collection chamber, and was sampled after 15, 30 and 60 minutes. The concentration of nicardipine remained as expected with no loss regardless of the location, time of sampling and exposure to light. All results are presented as mean ± standard deviation. Dashed line represents the expected nicardipine concentration. ns - non-significant.

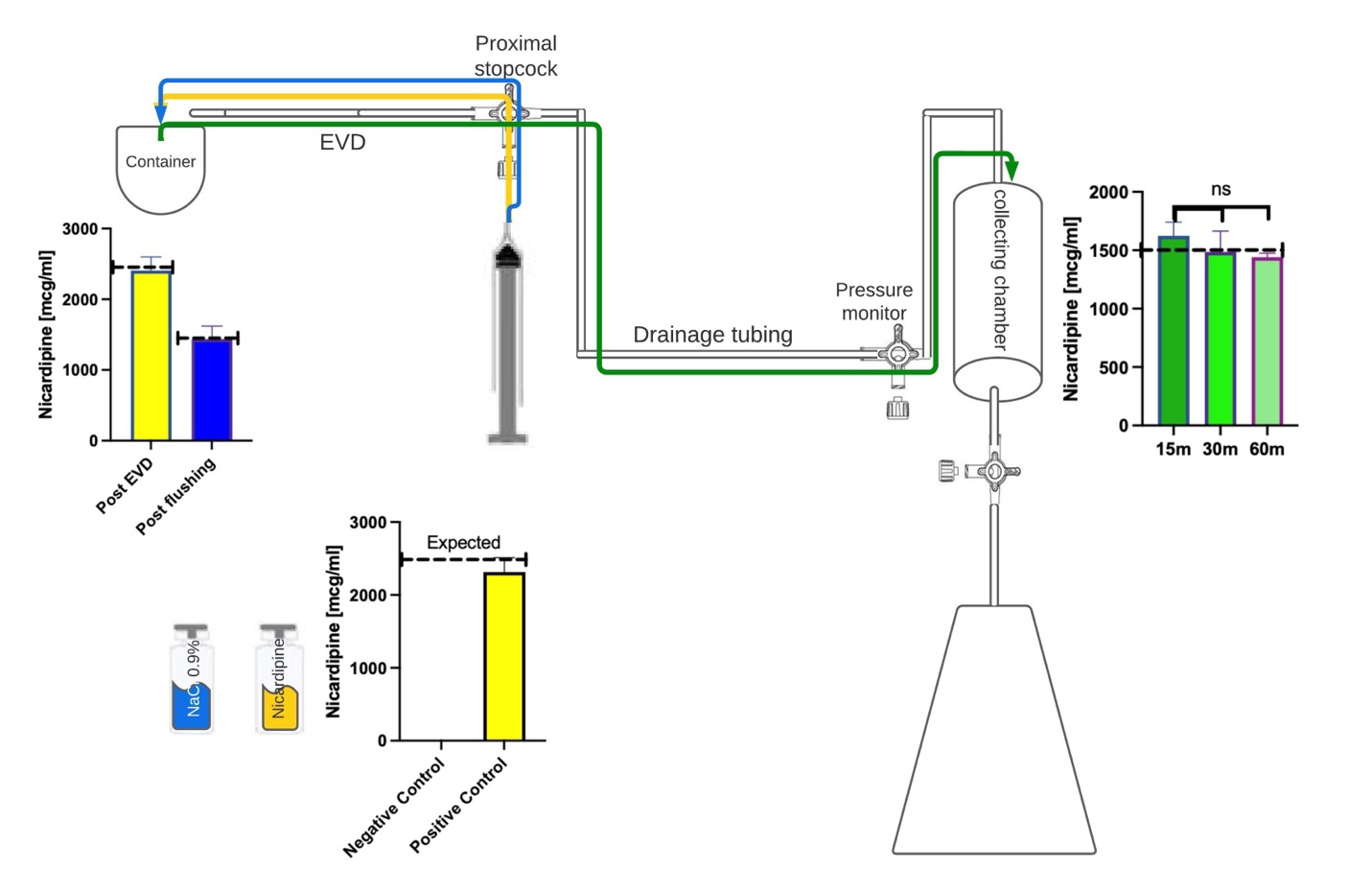

**Supplementary Figure S2.** Goodness-of-fit plots for the final PK model of IT nicardipine in SAH patients.

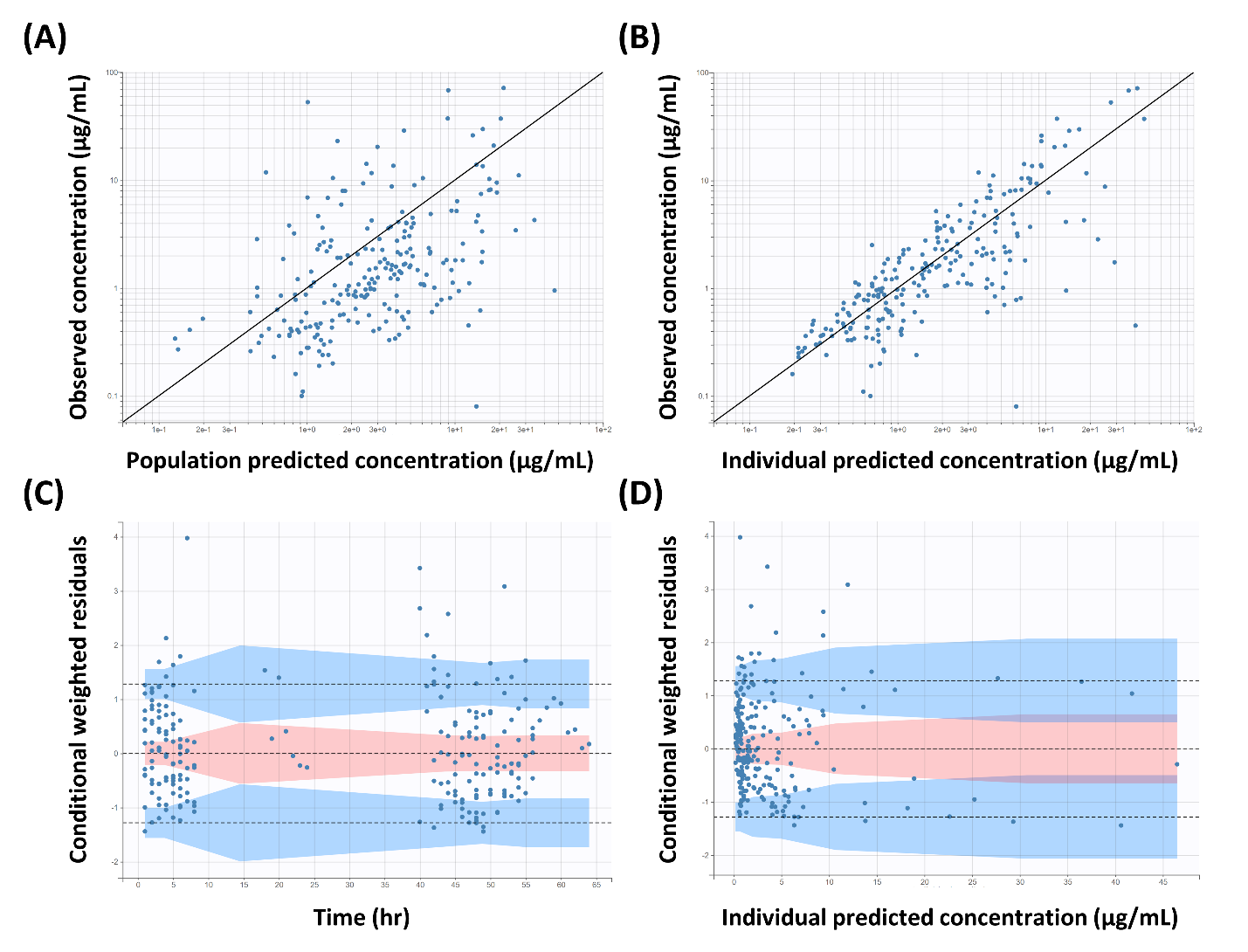

**Supplementary Figure S3.** Visual predictive check (VPC) for the final PK model of nicardipine in SAH patients. All CSF concentration observations and predictions were adjusted using prediction correction.^28^ The lines denote the 10^th^, 50^th^, and 90^th^ percentiles of the observed data, while prediction intervals are depicted as the pink (the 50^th^ percentile) and blue areas (the 10^th^ and 90^th^ percentiles).

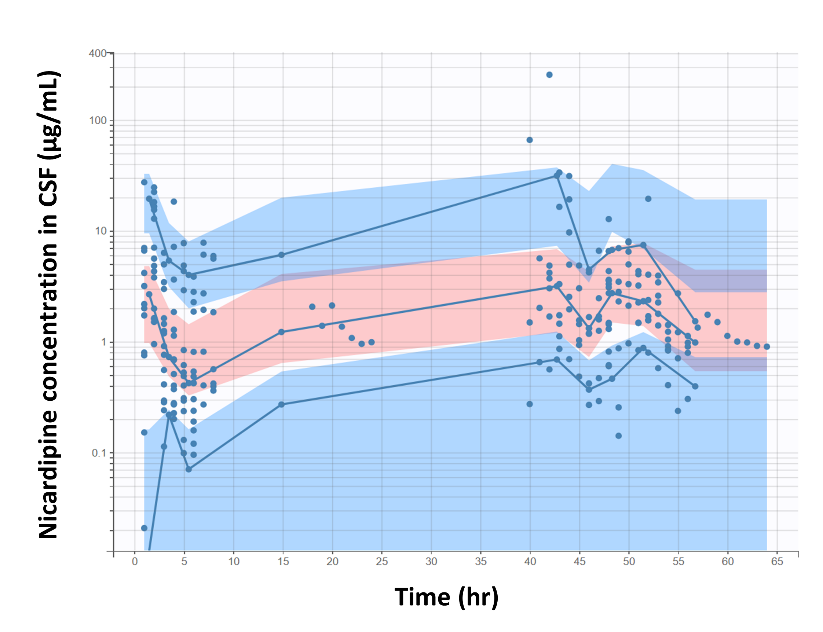
